# Evaluate the risk of resumption of business for the states of New York, New Jersey and Connecticut via a pre-symptomatic and asymptomatic transmission model of COVID-19

**DOI:** 10.1101/2020.05.16.20103747

**Authors:** Ting Tian, Jianbin Tan, Yukang Jiang, Xueqin Wang, Heping Zhang

## Abstract

The United States has the highest number of confirmed cases of COVID-19 in the world. The early hot spot states were New York, New Jersey, and Connecticut. The workforce in these states was required to work from home except for essential services. It was necessary to evaluate an appropriate date for resumption of business since the premature reopening of the economy would lead to a broader spread of COVID-19, while the opposite situation would cause greater loss of economy. To reflect the real-time risk of the spread of COVID-19, it was crucial to evaluate the population of infected individuals before or never being confirmed due to the pre-symptomatic and asymptomatic transmissions of COVID-19. To this end, we proposed an epidemic model and applied it to evaluate the real-time risk of epidemic for the states of New York, New Jersey, and Connecticut. We used California as the benchmark state because California began a phased reopening on May 8, 2020. The dates on which the estimated numbers of unidentified infectious individuals per 100,000 for states of New York, New Jersey, and Connecticut were close to those in California on May 8, 2020, were June 1, 22, and 22, 2020, respectively. By the practice in California, New York, New Jersey, and Connecticut might consider reopening their business. Meanwhile, according to our simulation models, to prevent the resurgence of infections after reopening the economy, it would be crucial to maintain sufficient measures to limit the social distance after the resumption of businesses. This precaution turned out to be critical as the situation in California quickly deteriorated after our analysis was completed and its interventions after the reopening of business were not as effective as those in New York, New Jersey, and Connecticut.

## 1 Introduction

The outbreak of novel coronavirus disease (COVID-19) has spread over 200 countries since December 2019 (National Health Commission of the People’s Republic of China, 2020). It is unprecedented to have over 7 million cumulative confirmed cases of COVID-19 worldwide at the beginning of June, 2020 (World Health Organization, 2020c). The “battle” against COVID-19 in China has provided experience and likely outcomes of certain interventions to the ongoing hard-hit areas. As a novel and acute infectious disease, the transmission mechanisms of COVID-19 were unknown at the early stage of the epidemic, and the Chinese government implemented relatively strict non-pharmaceutical interventions in the hot spot areas, where the public transportation was suspended within and outside of the cities in Hubei province since January 23, 2020 (Chinese Center for disease control and prevention, 2020). All nationwide residents were recommended to stay at home except for essential needs. The holiday season of the Chinese Spring Festival had been prolonged until late February when essential services were recommenced operating gradually outside Hubei province (The State Council, The People’s Republic of China, 2020b). In April 2020, a comprehensive resumption of business started in China (The State Council, The People’s Republic of China, 2020a).

In late January 2020, the United States began reporting confirmed cases of COVID-19 (Holshue et. al., 2020). There were over 1,000 cumulative confirmed cases on March 13, 2020 (World Health Organization, 2020a), when the White House declared a national emergency concerning COVID-19 outbreak (The White House, 2020b) and issued a “call to action” coronavirus guidelines on March 16, 2020 (The White House, 2020a). The United States has become the most severe country of COVID-19 with 366,346, 154,154, 40,468, and 94,743 cumulative confirmed cases in the states of New York, New Jersey, Connecticut, and California by May 24, 2020 (The Center for Systems Science and Engineering (CSSE) at Johns Hopkins University, 2020), respectively. Making things worse, New York and California are the top two states that contribute to the real gross domestic product (GDP) in the United States (Figure 1) (The United States Census Bureau, 2020).

**Figure 1:**
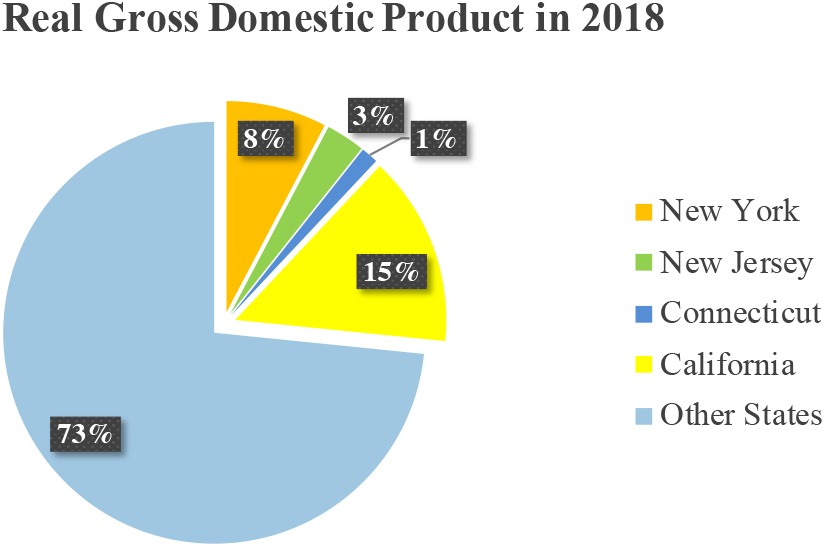
The percentages of Gross Domestic Product of New York, New Jersey, Connecticut, and California in 2018.

The state of New York reported the first confirmed case of COVID-19 on March 1, 2020, and proclaimed an executive order on March 16, 2020, including reducing half of the local government workforce, allowing the statewide nonessential workforce to work from home starting on March 17, 2020, and closing all schools starting on March 18, 2020 (State of New York Governor Andrew Cuomo, 2020g). Due to the rapidly increasing number of additional cases of COVID-19 in the state, the governor announced an aggressive policy of “New York State on PAUSE” on March 20, 2020 (State of New York Governor Andrew Cuomo, 2020c), and required all people in the state to wear masks or face covering in public since April 15, 2020 (State of New York Governor Andrew Cuomo, 2020f). On May 1, 2020, all statewide K-12 schools and college facilities continued to close for the remaining academic year (State of New York Governor Andrew Cuomo, 2020a). The guide of the “NY Forward Reopening” Plan was available on May 11, 2020, and three regions reopened businesses for phase one on May 15, 2020 (State of New York Governor Andrew Cuomo, 2020b). By May 24, 2020, seven regions reopened for businesses (State of New York Governor Andrew Cuomo, 2020e).

The state of New Jersey reported the first positive case of COVID-19 on March 4, 2020 (State of New Jersey Governor Phil Murphy, 2020a). The governor of New Jersey recommended the cancellation of statewide public gatherings over 250 individuals from March 12, 2020, suspended visiting state prisons and statewide halfway houses effective on March 14, 2020, and closed restaurants, bars, movie theaters, gyms, casinos from March 16, 2020 (State of New Jersey Governor Phil Murphy, 2020e). The governor announced an order including the statewide stay at home and closure of statewide non-essential retail industries on March 21, 2020 (State of New Jersey Governor Phil Murphy, 2020c). State parks and golf courses reopened on May 2, 2020 (State of New Jersey Governor Phil Murphy, 2020g). Among other activities, construction and non-essential retail provisions were allowed to resume on May 18, 2020 (State of New Jersey Governor Phil Murphy, 2020f). In-person sales were authorized to reopen at the car, motorcycle, and boat dealerships and bike shops by appointment only and with social distancing measure on May 20,2020 (State of New Jersey Governor Phil Murphy, 2020b).

The state of Connecticut reported the first positive case on March 8, 2020 (State of Connecticut Governor Ned Lamont, 2020c). New York, New Jersey, and Connecticut announced measures to slow the spread of COVID-19 throughout the tri-state area on March 16, 2020 (State of Connecticut Governor Ned Lamont, 2020f). The governor signed an executive order for businesses and residents “stay safe, stay home” on March 20, 2020 (State of Connecticut Governor Ned Lamont, 2020e). Statewide K-12 schools were announced to remain closed for the rest of the academic year on May 5, 2020 (State of Connecticut Governor Ned Lamont, 2020b). The restaurants, offices, retail stores, outdoor museum, and zoos were authorized to reopen as the first phase on May 20, 2020 (State of Connecticut Governor Ned Lamont, 2020d) and hair salons and barbershops were aligned to reopen in early June (State of Connecticut Governor Ned Lamont, 2020a). New York, New Jersey, and Connecticut signed a multi-state agreement on the reopening of beaches on May 22, 2020 (State of New Jersey Governor Phil Murphy, 2020d).

The state of California reported two positive cases of COVID-19 on January 26, 2020 (Office of Public Affairs, 2020d). One month later, there was the first possible case of local transmission of COIVD-19 in California (Office of Public Affairs, 2020a). The “stay home except for essential needs” order was issued on March 19, 2020, where all individuals were required to stay at home except for the workforce in 16 critical infrastructure sectors (Office of Public Affairs, 2020b). On May 7, 2020, the state public health officer determined that the entire state gradually moved to Stage 2 of California’s Pandemic Resilience Roadmap, i.e., reopening the lower-risk workplaces and other spaces (Office of Public Affairs, 2020c). Therefore, California state had begun a phased reopening on May 8, 2020.

Due to the epidemic of COVID-19, the entire social system was slowed down and the unemployment in total nonfarm dramatically increased (Figure 8 in the supplement). The unemployment rates were 14.7% and 13.3% in April and May 2020 (U.S. Department of Labor, 2020). Thus, the time to restore the economy in the United States, especially for the states of New York, New Jersey, Connecticut, had been the most significant and consequential decision for the president of the United States and the governors of the states. For people to return to work, safety is a key concern. On April 13, 2020, seven states, including the states of New York, New Jersey, and Connecticut, joined an effort to form a multi-state council to reopen the economy while combating COVID-19 (State of New York Governor Andrew Cuomo, 2020d). The timelines of interventions in New York, New Jersey, Connecticut, and California are displayed in Figure 2.

**Figure 2:**
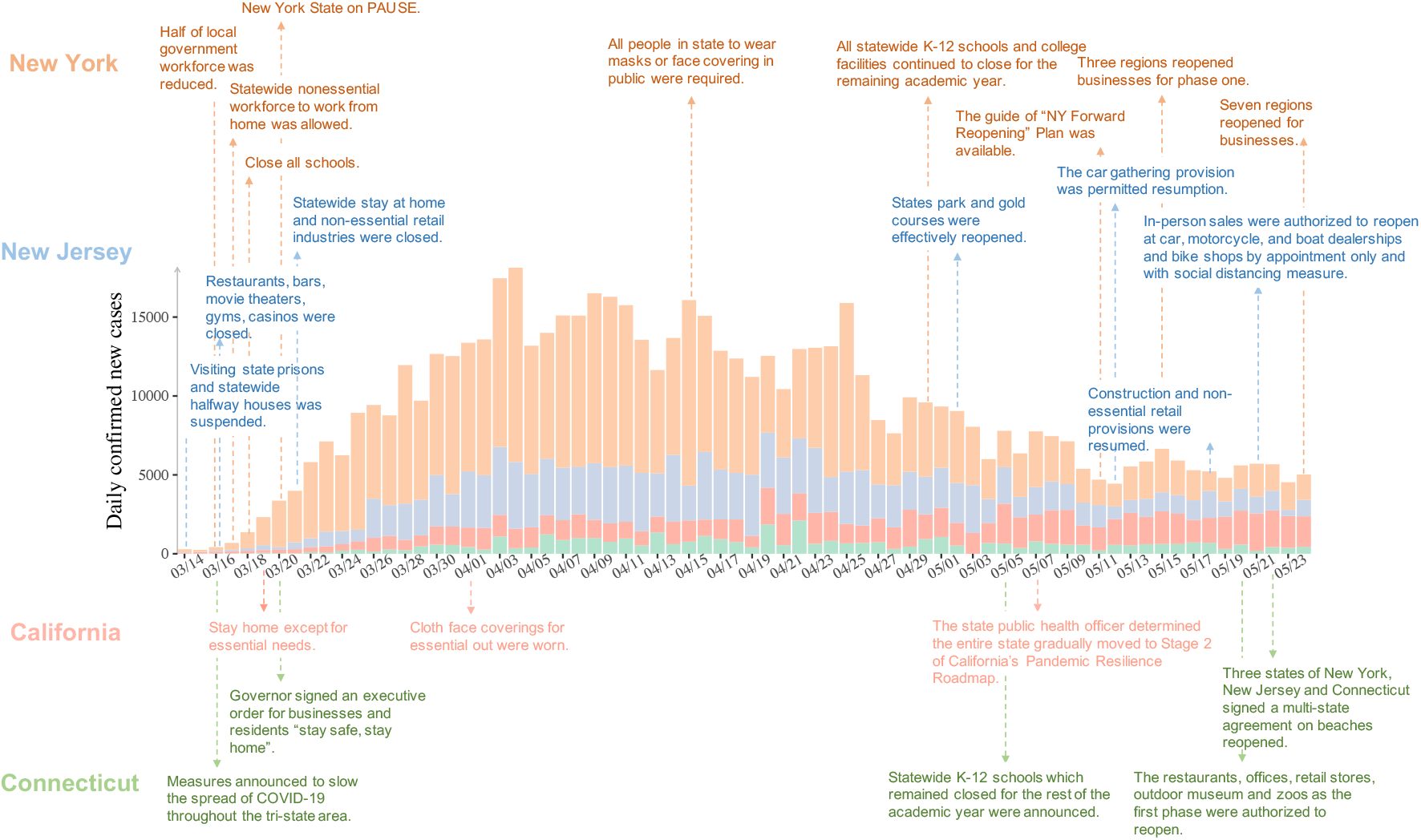
The timeline of interventions against COVID-19 in the states of New York, New Jersey, Connecticut, and California.

## 2 Methods

### 2.1 Data collection

This work began in April and initially completed in May 2020 when there were no widespread pretests of social injustice in the United States (Minnesota Daily, 2020). To emphasize the effectiveness of our model for evaluating the real-time risk of the epidemic, we chose to focus on the data before June 2020. This strategy also allowed us the opportunity to contrast our prediction and recommendations to what took place after June 2020.

We collected the epidemic data from March 13, 2020 when the national emergency concerning COVID-19 was proclaimed to May 24, 2020 in New York, New Jersey, Connecticut, and California. The data were made available by the New York Times (The New York Times, 2020).

### 2.2 Bayesian modelling of epidemic

Based on a WHO report, the transmission of COVID-19 could be caused by the individuals infected with the virus before significant symptoms developed (World Health Organization, 2020b), or even the carriers who did not develop symptoms. Pre-symptomatic transmission and asymptomatic transmission interfere with our ability to understand the real magnitude of COVID-19 because of the lag from the time of catching the virus to the time of being confirmed by testing. To overcome this issue, we divided a concerned population into four compartments: susceptible (*S*), unidentified infectious (*I*), self-healing without being confirmed (*H*), and confirmed cases (*C*).

- The susceptible (S) individuals have no immunity to the disease and are the majority of the population at an early stage of the epidemic.
- Unidentified infectious (*I*) individuals are infectious but not confirmed individuals, and can be divided into two groups: those who would eventually develop symptoms and the others who would not develop symptoms called asymptomatic carriers. We assumed that individuals in compartment *I* would move into either group *H* or *C* eventually.
- Self-healing individuals without being confirmed (*H*) are assumed to be no longer infectious and resistant to COVID-19.
- Confirmed cases (*C*) include two groups of individuals: patients in the hospital and asymptomatic carriers who are supposed to stay at home, and unable to transmit the disease.

We introduced Susceptible (*S*)-Unidentified infectious (*I*)-Self-healing without being confirmed (ii)-Confirmed cases (*C*) (SIHC) model to accommodate the four compartments above. Figure 3 presents the flowchart among them.

**Figure 3:**
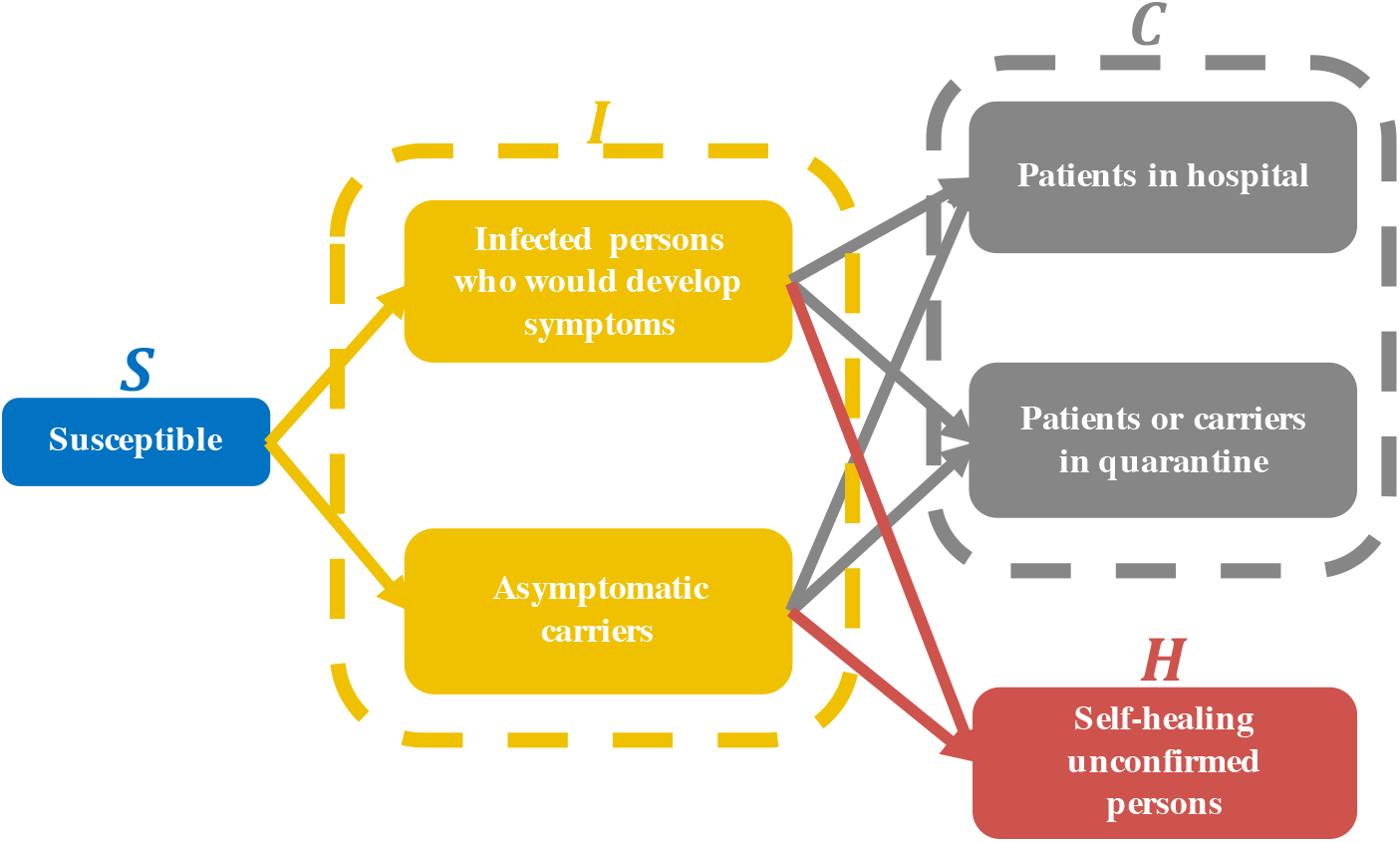
The transition diagram among compartments *S, I, H*, and *C*.

We assumed the following dynamic system in terms of the numbers of individuals in compartments *S*, *I, H*, and *C* at time t, denoted by *S*(*t*), *I*(*t*), *H*(*t*), and *C*(*t*), respectively,

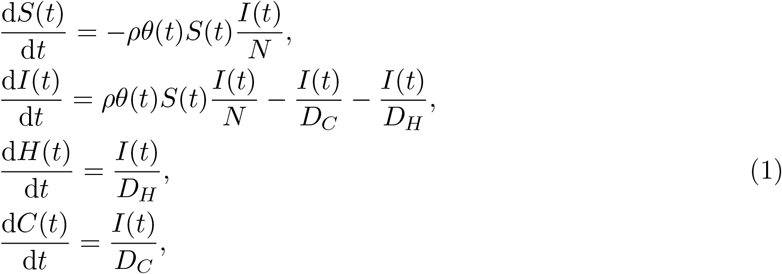

where *ρ* is the transmissiblility and *θ*(*t*) is the average contact number per person at time *t* (Blackwood and Childs, 2018), which was assumed to be time-varying; *D_H_* is the average duration from catching the virus to self-healing without being confirmed; *D_C_* is the average duration from catching the virus to be confirmed by testing; and *N* is the total number of population.

Let *α*(*t*) = *ρθ*(*t*)*. θ*(*t*) can be controlled by policy interventions. Typically, it is a constant at an early stage of the epidemic and decreases as the interventions are implemented until the interventions take full effect when it reaches or nears the lowest level. Based on this point of view, we further assumed that *α*(*t*) was a monotonically decreasing curve (Tan et al., 2020) with four parameters *α*_0_, *d, m*, and *η*

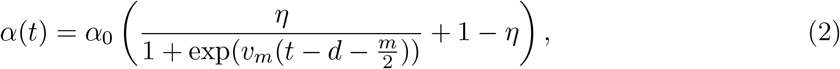

where *α*_0_ denotes the maximum of *α*(*t*) at an early stage of the outbreak, *d* is the time that takes for the control measures to start their effects and for *α*(*t*) to start declining, (1 − *η*) is the ratio of *α*_0_ to the minimum of *α*(*t*), which is *α*_0_(1 − *η*). *υ_m_* was chosen as 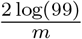 so that *α*(*t*) first reaches the minimum at *d* + *m*. Figure 4 illustrates the shape of *α*(*t*).

**Figure 4:**
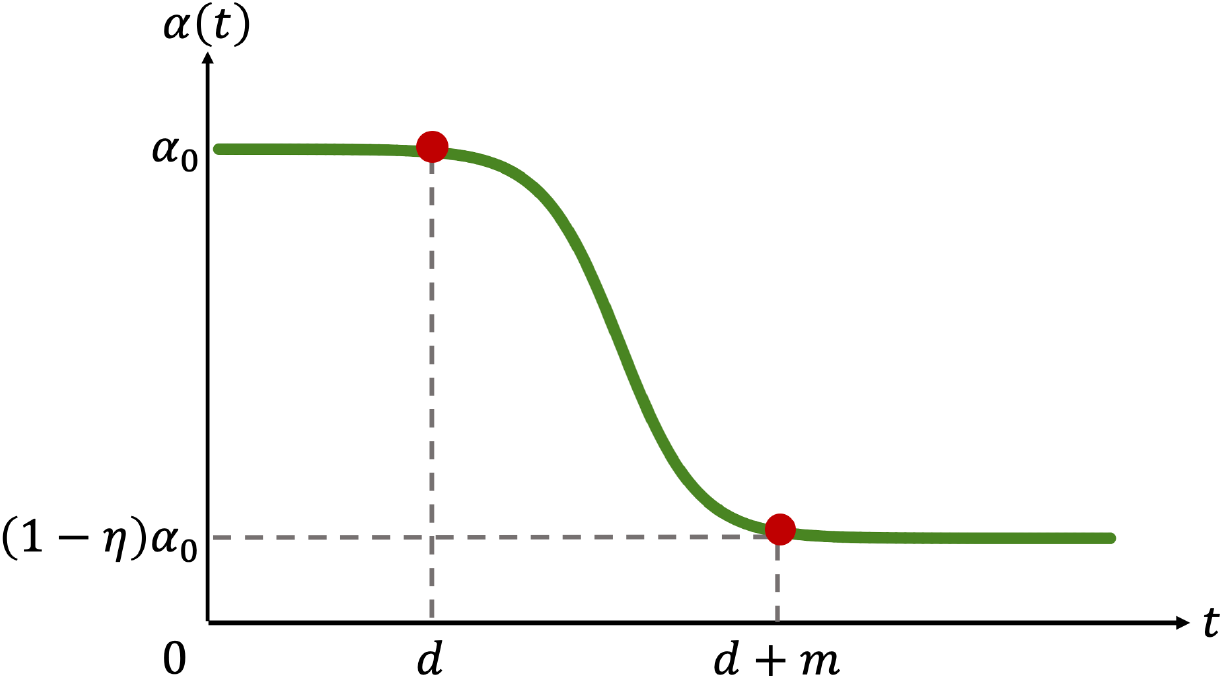
An illustration of *α*(*t*).

Next, we derived the time-varying reproduction number *R_t_* (Anderson et al., 1992; Jones, 2007) by our SIHC model:

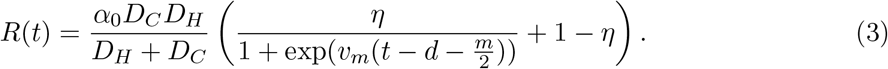

Let Ω = (*α*_0_, *η, m, D_C_*) and Θ = (Ω, *d,D_H_*), where *d* and *D_H_* were given and the others needed to be estimated. Note that

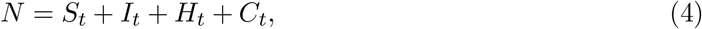

and *Z_t_* = (*I_t_*, *H_t_*, *C_t_*) was assumed to be a latent Markov process:

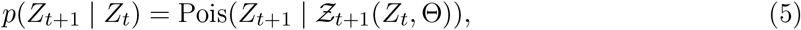

where *Ƶ*_t+1_ (*Z_t_*, Θ) was the evolving operator to determine the values of *I*, *H* and *C* at time (*t* + 1) given the deterministic dynamic system (1) with the initial value of (*N* − *I_t_* − *H_t_* − *C_t_*, *I_t_*, *H_t_, C_t_*) and parameters Θ. Pois(·|λ) is the mass of a multi-dimensional independent Poisson distribution with mean vector λ. Similarly, we defined 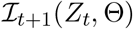, 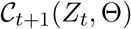, 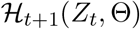, as the components of 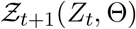. Since *N*, *S_t_* ≫ *I_t_*, *H_t_*, *C_t_*, the conditional independent Poisson likelihood of (*I_t_*_+1_, *H_t_*_+1_, *C_t_*_+1_) is the Poisson approximation for the multinomial likelihood of (*S_t_*_+1_, *I_t_*_+1_, *H_t_*_+1_, *C_t_*_+1_), whose incident rate is 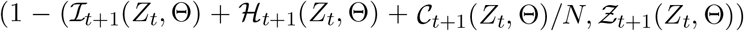 and the total number is *N* (Tan et al., 2020). Notice that Ƶ_t+1_(*Z_t_*, Θ) has no closed form, we used **ode**() function implemented by R package **deSolve** (Soetaert et al., 2010) to solve the given system of ordinary differential equations.

Given the observable data *C*_1:_*_T_* and *N*, where *T* was the time period of observation, we set *H*_1_ = 0, which was reasonable for the early stage of the outbreak. The numbers of individuals in compartments *I*_1:_*_T_* and *H*_2:_*_T_* were treated as the latent variables. For simplicity, we assumed that *α*(*t*) ≡ *α*(*k*), if *k* ≤ *t* < *k* + 1. Moreover, we assumed that 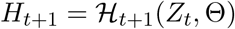 a.s.; namely, *H_t_*_+1_ depended completely on *I_t_*, *H_t_* and *D_H_*. In fact, our empirical observation suggested that this simplification did not substantially alter the results, while assuming 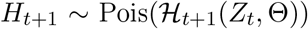 would have substantially increased the computational burden for the sampling procedure. We employed a Bayesian procedure in our parameter estimation and future prediction. The posterior distribution of the parameters was (Bolstad and Curran, 2016):

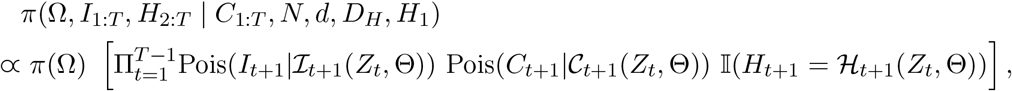

where π(·) represents the prior distribution of corresponding parameter and π(·|*) represents the posterior distribution of corresponding parameter given the observed data “*”. Similarly, we can use the posterior distribution of *Z_s_*: π(*Z_s_*|*C*_1:_*_T_*, *N, d*, *D_H_, H*_1_) (*s* > *T*), to predict the spread of infectious disease.

For prior distributions, notice that *D_C_* was governed by the mean of the incubation period. Based on the related literature on the incubation period of COVID-19 (Lauer et al., 2020), we chose an informative prior: the log-normal distribution with the log-mean of log(5.1) as the prior distribution of *D_C_*. Similarly, we chose the log-normal distribution with the log-mean of log(30) as the prior distribution of *m*. In other words, the interventions were assumed to take the full effect after one month. To enhance the influence of prior information, we assumed the log-standard deviation for both *D_C_* and *m* is log(1.05). The priors of the remaining parameters were chosen to be non-informative or flat priors, i.e., *π*(*α*_0_), *π*(*η*) ∝ 1.

For the fixed parameters *d*, and *D_H_*, recall that *d* was the waiting time for interventions to begin. We chose *d* as the start of implementing certain interventions, i.e, *d* = 8 for New York (see “New York State on PAUSE” in Figure 2), *d* = 9 for New Jersey (see “statewide stay at home” in Figure 2), *d* = 8 for Connecticut (see “stay safe, stay home” in Figure 2), and *d* = 7 for California (see “stay home except for essential needs” in Figure 2). *D_H_* was assumed to be 9.5 according to the clinical study of asymptomatic cases (Hu et al., 2020).

Since *π*(Ω, *I*_1:T_, *H*_2:_*_T_*|*C*_1:_*_T_*, *N*, *d*, *D_H_*, *H*_1_) has no closed form, we used Markov Chain Monte Carlo (MCMC) (Ghosh et al., 2006; Andrieu et al., 2003; Chib and Greenberg, 1995; Soubeyrand, 2016) to approximate posterior distributions of parameters Ω and latent numbers of *I* and *H* at each iteration. The Supplement provides the details of the sampling algorithm. The data and the R files relevant to the analysis in this study are available at https://github.com/tingT0929/Resumption-of-business. The point estimates of Ω, *I*_1:_*_T_*, *H*_2:_*_T_* and the prediction of spread were the medians of the posterior distribution while 95% credible intervals were constructed with 2.5% and 97.5% quantiles.

## 3 Data analysis

### 3.1 Model estimation

We used publicly available data to simulate the possible outcomes of the outbreak of COVID-19 by varying the dates when the business reopens. Table 1 presented the parameter estimates. These estimates were then used in our simulation models for potential second waves of COVID-19 while we assessed the risk for people to return to work. The data after our models were built and used to assess the validity of our simulation models.

**Table 1:** The estimated parameters and their corresponding confidence intervals.

|  | New York | New Jersey | Connecticut | California |
| --- | --- | --- | --- | --- |
| $\hat{\alpha}_0$ | 0.67 (0.63, 0.69) | 0.57 (0.56, 0.58) | 0.50 (0.47, 0.52) | 0.45 (0.43, 0.46) |
| $\hat{\eta}$ | 0.33 (0.31, 0.34) | 0.44 (0.43, 0.46) | 0.42 (0.39, 0.44) | 0.29 (0.25, 0.31) |
| $\hat{m}$ | 16.84 (15.86, 17.36) | 18.66 (18.08, 19.38) | 30.39 (28.28, 31.76) | 26.74 (24.54, 27.84) |
| $\hat{D}_C$ | 2.22 (2.11, 2.35) | 3.15 (3.05, 3.24) | 3.47 (3.27, 3.68) | 3.14 (2.96, 3.25) |

Briefly, the average relative error between our predicted and the observed numbers of cases from May 25 to June 6, 2020 was computed from

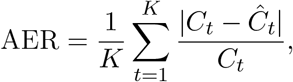

where *K* is the length of prediction time. The AERs were 0.34%, 0.45%, 0.59%, 3.56% in New York, New Jersey, Connecticut, and California, respectively. These low levels of errors suggested accurate prediction from our models. The estimated trends for the four states are given in Figure 9 in the Supplement.

The estimated rates of infected individuals without being confirmed on May 24, 2020 (i.e. the estimated 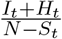 for time t to be May 24, 2020) were 22.82% with 95% CI (21.86%, 24.68%), 32.38% with 95% CI (31.36%, 33.09%), 34.95% with 95% CI (33.04%, 36.32%), 37.74% with 95% CI (35.72%, 38.53%), respectively, in the four states. It had been reported that the numbers of tests per 100,000 in New York, New Jersey, Connecticut, and California on May 24, 2020, were 9312, 7434, 6321, and 4396, respectively (Jin, 2020). It appeared that the rate of testing was inversely proportional to the rate of unidentified infected.

To visualize the importance of the unobserved number of unidentified infected individuals (*N* − *S_t_*), rather than the observed confirmed cases (*C_t_*), for the assessment of epidemic risk, we displayed the comparison between the estimated number of new daily infected individuals and observed new daily confirmed cases in Figure 5. Note that there were gaps between the number of new daily confirmed cases and new infected individuals in Figure 5 as a result of the pre-symptomatic and asymptomatic transmissions of COVID-19. This phenomenon was the lagging effect for new daily confirmed cases, reflecting the time interval from catching the virus to being confirmed by testing. Figure 5 indicated that we cannot ignore the lagging effect at an early stage of the epidemic, although it seemed to disappear over time.

**Figure 5:**
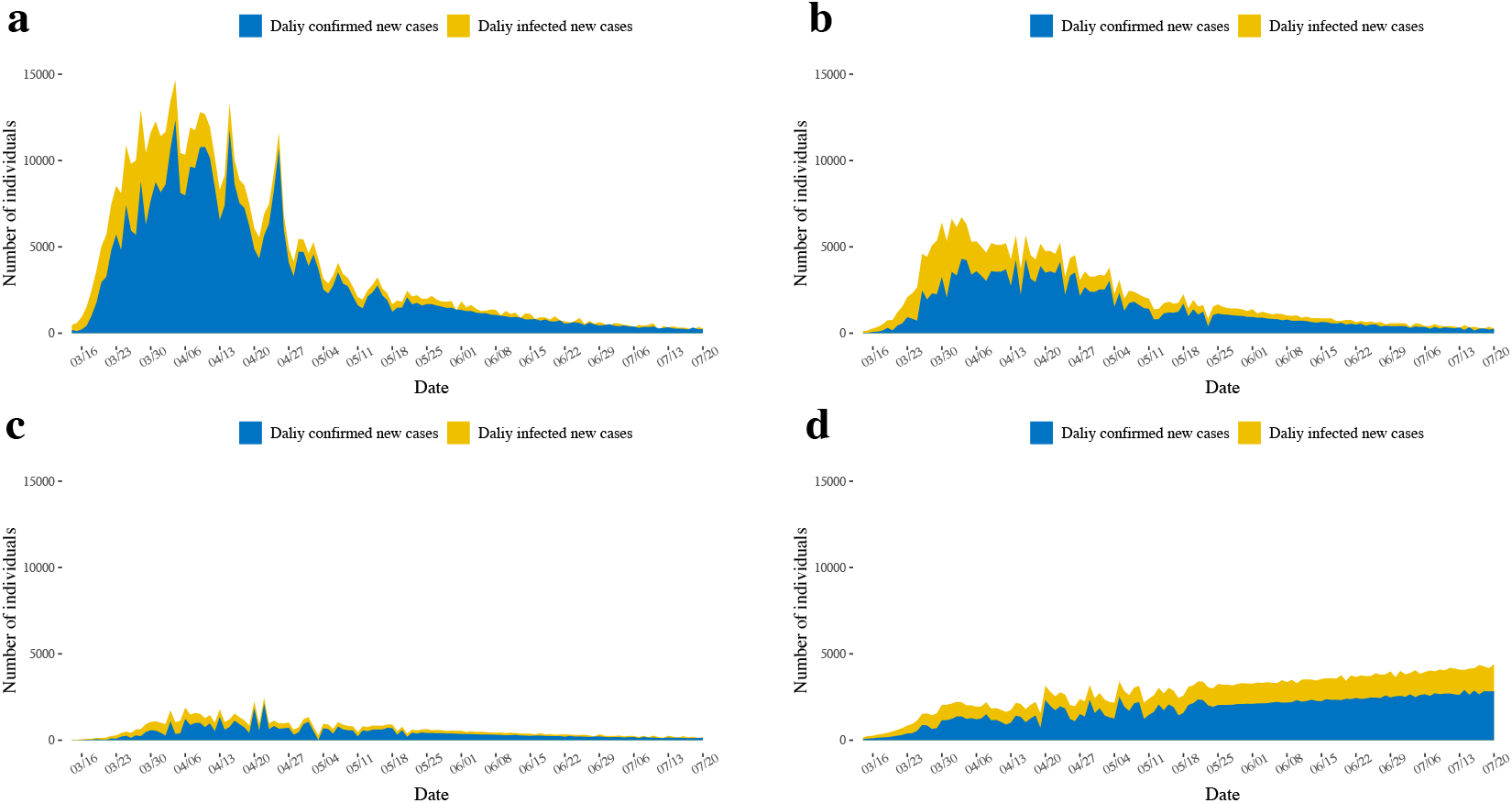
The magnitude of the outbreak of COVID-19 in New York (a), New Jersey (b), Connecticut (c), and California (d) from March 13 to July 20, 2020. The daily confirmed new cases from March 13 to May 24, 2020, were observed data, and the data from May 25 to July 20, 2020, were projected by our models.

### 3.2 Evaluate the risk for reopening the economy

To appreciate the potential risk of COVID-19, we considered the estimated numbers of the only transmissible compartment *I_t_* per 100,000 for New York, New Jersey, Connecticut, and California; see Figure 6. This figure indicated that the peak of unidentified infectious individuals in New York, New Jersey, and Connecticut had passed. So far, this remained to be the case.

**Figure 6:**
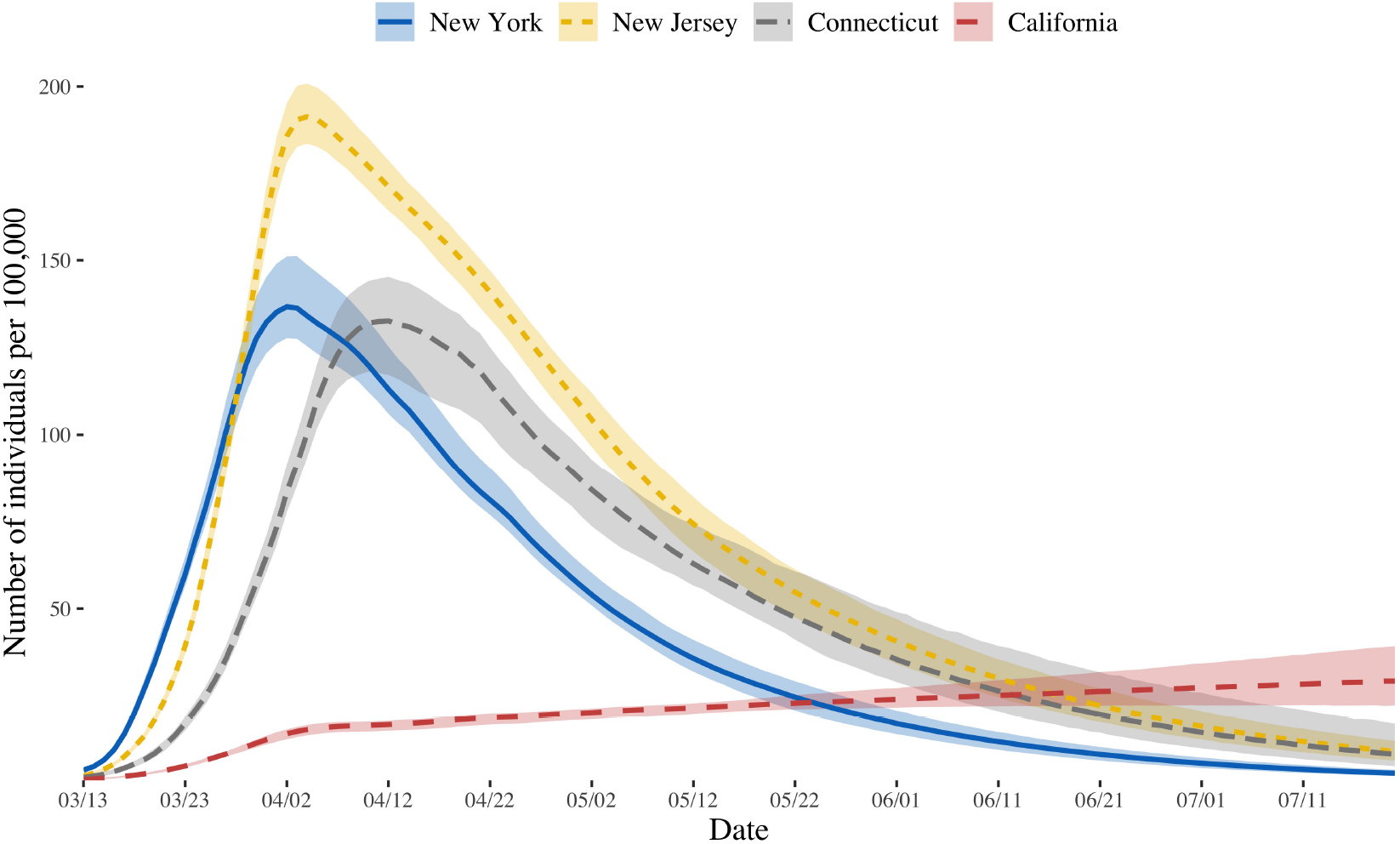
The trend of the numbers of unidentified infectious individuals per 100,000 for states of New York, New Jersey, Connecticut, and California from March 13 to July 20, 2020. The corresponding 95% credible intervals were represented in each state.

California reopened the lower-risk workplaces as Stage 2 on May 8, 2020. This decision of California was used as a reference in our consideration of resuming the business in New York, New Jersey, and Connecticut. Compared with California, the numbers of unidentified infected individuals (*I_t_*) in New York, New Jersey, and Connecticut were higher before May 8, 2020, but on clearly descending trajectories. However, while the number was low, the steady upward trajectory in California was concerning. This upward trajectory coupled with insufficient interventions might be the major cause for the resurgence of COVID-19 in California after the reopening.

To balance the risk of epidemic versus resuming business, we considered a few choices of Mondays in June 2020 as possible dates of reopening. We used our simulation models to predict the numbers of unidentified infectious individuals (*I_t_*) per 100,000 for New York, New Jersey, Connecticut, and California on June 1, June 8, June 15, June 22 and June 29, 2020. The results were given in Table 2. Common wisdom is that the risk is regarded as reasonably low for a population when the number of unidentified infectious persons per 100,000 is closed to 20. This guideline was used to form cross-state travel guidelines in the United States. This was then used as a rationale for a safe resumption of business.

**Table 2:** The estimated numbers of unidentified infectious individuals per 100,000 in the four states in the different Mondays.

|  | New York | New Jersey | Connecticut | California |
| --- | --- | --- | --- | --- |
| June 1 | 17.12 (14.07, 20.31) | 40.68 (34.18, 46.10) | 35.42 (28.93, 47.62) | 24.03 (21.63, 26.84) |
| June 8 | 13.29 (10.73, 15.93) | 32.88 (27.18, 37.80) | 28.93 (22.72, 40.57) | 24.80 (21.97, 28.21) |
| June 15 | 10.28 (8.22, 12.42) | 26.71 (21.65, 31.27) | 23.50 (17.77, 34.56) | 25.58 (22.07, 29.79) |
| June 22 | 7.97 (6.20, 9.77) | 21.52 (17.09, 25.76) | 19.08 (13.81, 29.66) | 26.33 (22.23, 31.23) |
| June 29 | 6.15 (4.73, 7.62) | 17.35 (13.41, 21.10) | 15.51 (10.69, 25.15) | 27.12 (22.29, 32.86) |

To appreciate the potential risk of resuming business on different Mondays, we simulated the possible second wave of infection after people returned to work. Unlike the first attack, we assumed that stringent interventions would be re-enforced one week after the business was reopened. We used the estimated 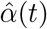 in Table 1 under “reopening” operation (6) in the Supplement as the parameters underlying the COVID-19 transmission.

The simulated results are given in Figure 7. Brief, for New York, the simulated numbers of cumulative confirmed cases per 100,000 on July 20 after the business was resumed on June 1, June 8, June 15, June 22 and June 29 were, respectively, 20.26%, 14.76%, 10.37%, 6.98%, and 4.37% higher than those if the business was not resumed. For New Jersey, the simulated numbers of cumulative confirmed cases per 100,000 on July 20 after the business was resumed on June 1, June 8, June 15, June 22 and June 29 were, respectively, 43.91%, 33.07%, 24.15%, 16.72% and 10.70% higher than those if the business was not resumed. For Connecticut, the simulated numbers of cumulative confirmed cases per 100,000 on July 20 after the business was resumed on June 1, June 8, June 15, June 22 and June 29 were, respectively, 37.18%, 28.08%, 20.45%, 14.29%, and 9.12%, higher than those if the business was not resumed.

**Figure 7:**
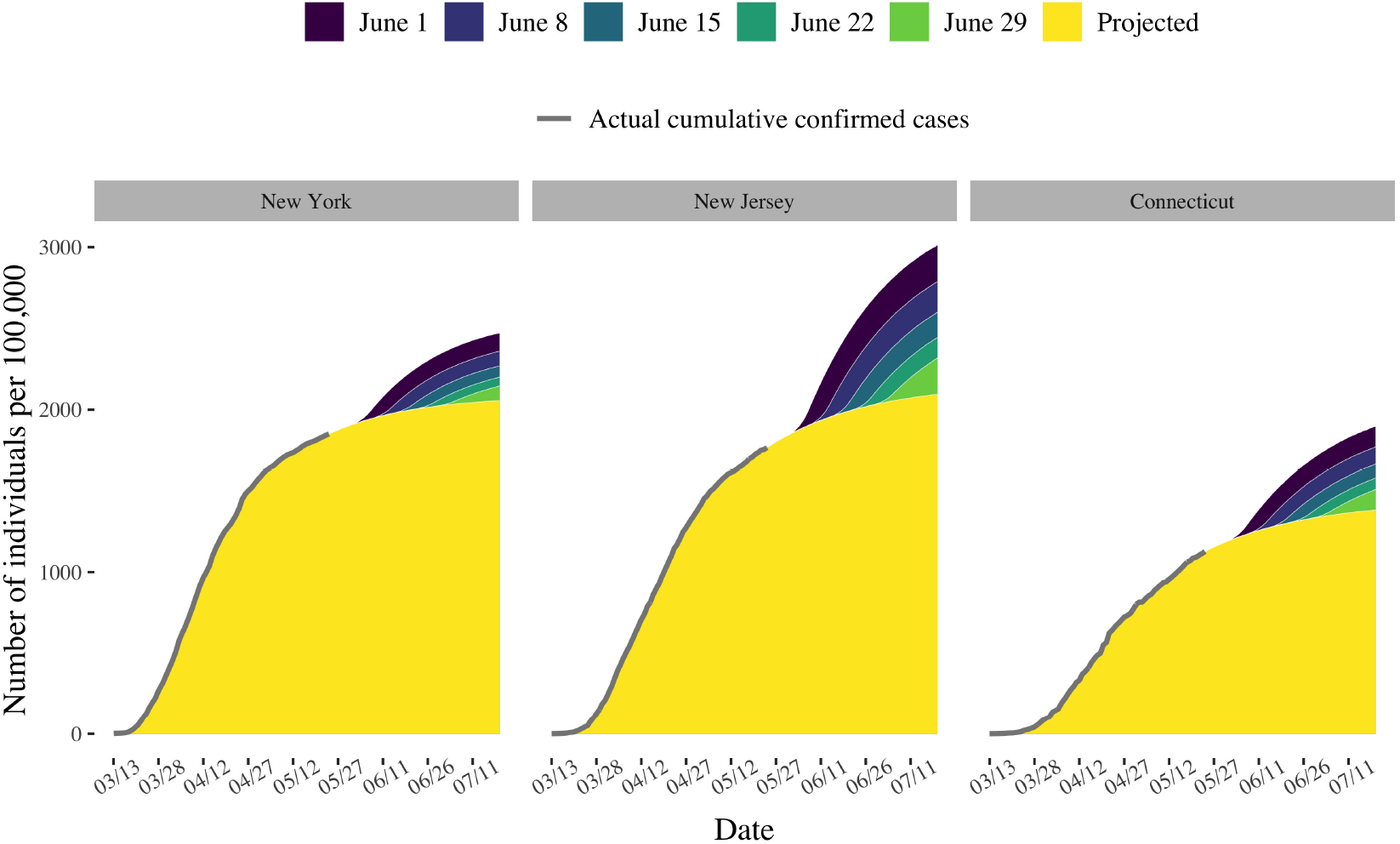
The possible outbreak of epidemic after people returned to work on different Mondays in New York, New Jersey, and Connecticut. The points represented the observed cumulative confirmed cases (per 100,000) in these states.

## 4 Discussion

The decision for the resumption of business is not only a public health issue but also an economic issue. What we focused on was the epidemiological feasibility of returning to work at an early date. There were obviously many other factors to consider (Centers for Disease Control and Prevention, 2020). To analyze the epidemic data of COVID-19 in New York, New Jersey, and Connecticut, we proposed an epidemic model by considering pre-symptomatic and asymptomatic transmissions of COVID-19. This model provided estimates for the numbers of new daily infected individuals and new confirmed cases. The higher number of unidentified infectious individuals, the higher risk for the resumption of business was expected. From Figure 7, we concluded that there were certain risks for the resumption of businesses on June 1, 2020, for New York, New Jersey, and Connecticut. If the governors of those states delayed the resumption of businesses for one week or more, the simulated magnitude of the second wave of the infection was much lower. However, the added benefit of delaying the reopening appeared less beyond one week and even more so after.

Because California began the process of reopening economy on May 8, 2020, we used the data in California at that time as a reference for reopening New York, New Jersey, and Connecticut. The dates when the estimated numbers of unidentified infectious individuals per 100,000 for states of New York, New Jersey, and Connecticut were close to those in California on May 8, 2020 were June 1, 22, and 22, 2020, respectively. By following the practice in California, New York, New Jersey, and Connecticut might consider reopening their business on June 1, 22, and 22, 2020, respectively. Moreover, we noted that the trajectory in California was clearly in a wrong direction as opposed to the descending trajectories New York, New Jersey, and Connecticut. While the three east coast states have become the lowest risk states, California has been suffering from a resurgence, underscoring the importance of maintaining public health practice after reopening business.

## Data Availability

This work began in April and initially completed in May 2020 when there were no widespread pretests of social injustice in the United States. To emphasize the effectiveness of our model for evaluating the real-time risk of the epidemic, we chose to focus on the data before June 2020. This strategy also allowed us the opportunity to contrast our prediction and recommendations to what took place after June 2020. 
We collected the epidemic data from March 13, 2020 when the national emergency concerning COVID-19 was proclaimed to May 24, 2020 in New York, New Jersey, Connecticut, and California. The data were made available by the New York Times.

https://covid-19.direct/state

https://www.mndaily.com/article/2020/06/pf-the-george-floyd-protests-a-visual-timeline

https://www.nytimes.com/interactive/2020/us/coronavirus-us-cases.html

## Supplement

**Figure 8:**
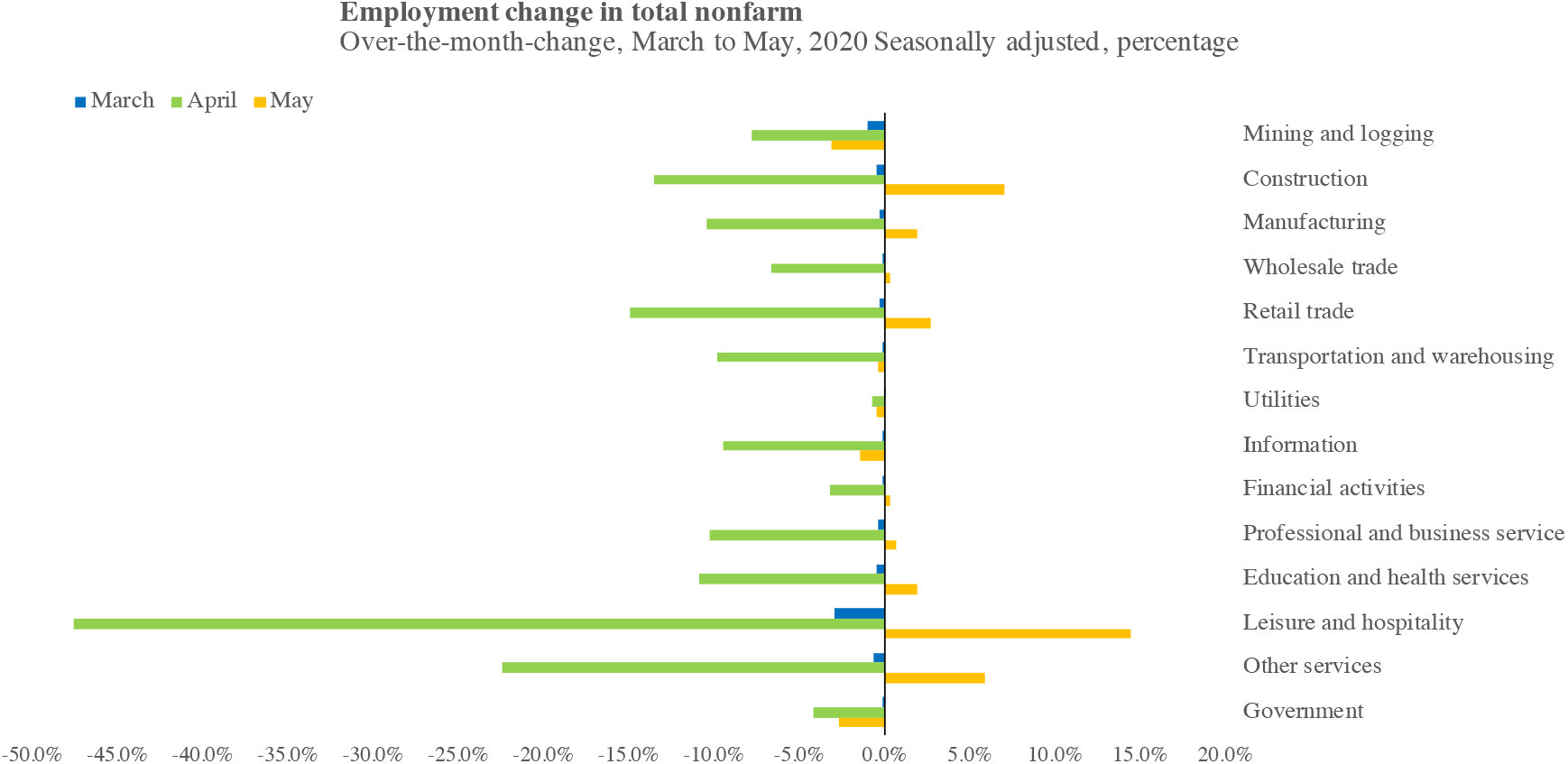
The over-the-month-change of employment in the total nonfarm sectors in the United States.

### Bayesian computation

Let 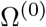, 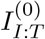, 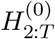 be the initial samples and Ω^(^*^k^*^)^, 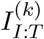 and 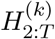 be the *k*-th samples. N_[0,∞]_(*μ*, *σ*^2^) represents the truncated normal distribution with mean *μ* and variance *σ*^2^ and logN(*μ*, *σ*^2^) represents the log-normal distribution with log-mean log(*μ*) and log-variance log(*σ*^2^). Algorithms 1, 2, and 3 provide detailed updating steps.

#### Algorithm 1 Update Ω

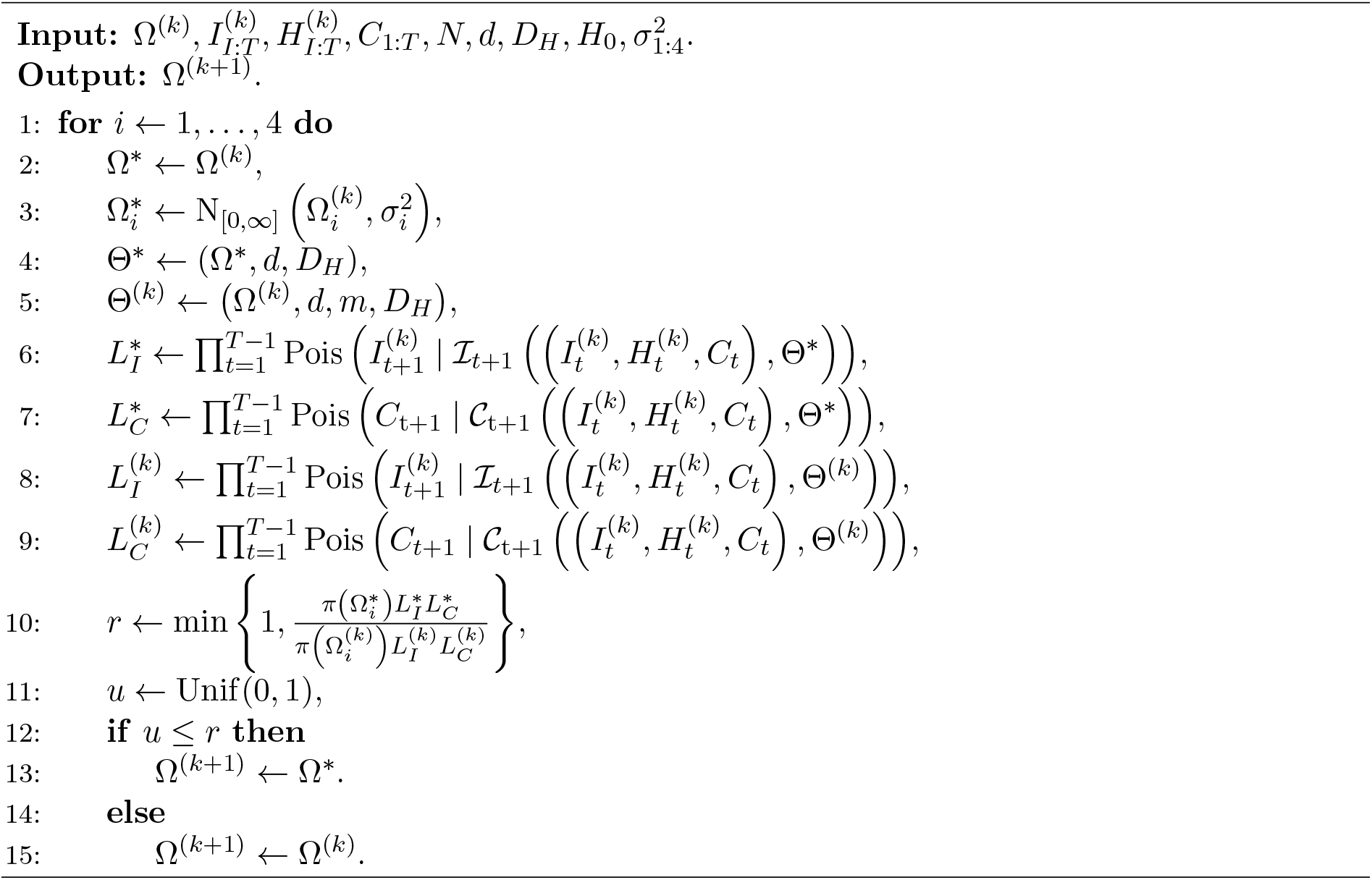

For sampling *I*_1_, notice that *S*_1_ ≈ *N*, which implied that

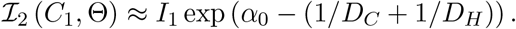

Then the full conditional distribution of *I*_1_ was approximated with

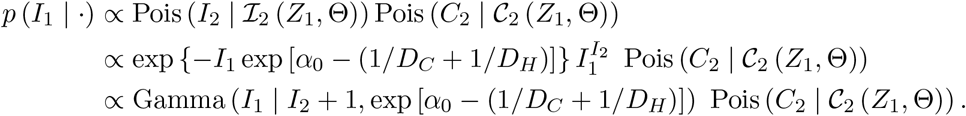

We use Gamma (*I*_2_ + 1, exp [*α*_0_ − (1/*D_C_* + 1/*D_H_*)]) as the proposal distribution of *I*_1_ in the Metropolis-Hastings algorithm.

#### Algorithm 2 Update *I*_1_

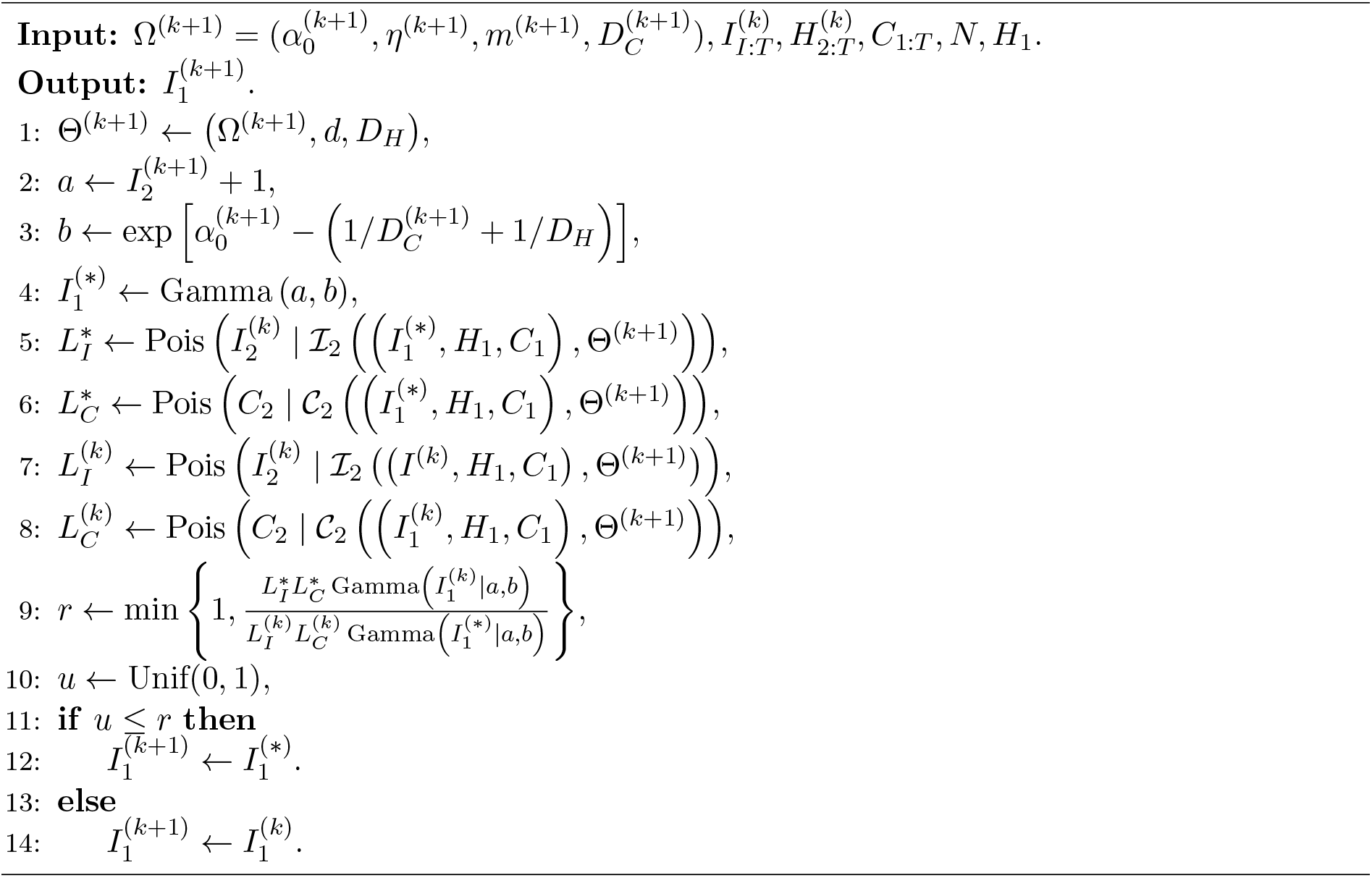

Moreover, there are two strategies to sample *I*_2:T_ and *H*_2:_*_T_*: sequential sampling (sequentially sample from *I_t_* and *H_t_* in each Metropolis-Hastings step or from the combined sample *I*_2:T_) and group sampling (sample from *H*_2:_*_T_*, and accept the samples in one Metropolis-Hastings step). The group sampling is high dimensional if *T* is large. Hence, moving from one iteration of MetropolisHastings algorithm to the next is computationally intensive. The sequential sampling requires more time for Markov chain to converge. We utilized the mixture of MCMC kernels Andrieu et al. (2003) to combine these two sampling strategies to balance the trade-off between the acceptance rate and the convergence time of MCMC.

#### Algorithm 3 Update *I*_2:T_, *H*_2:_*_T_*

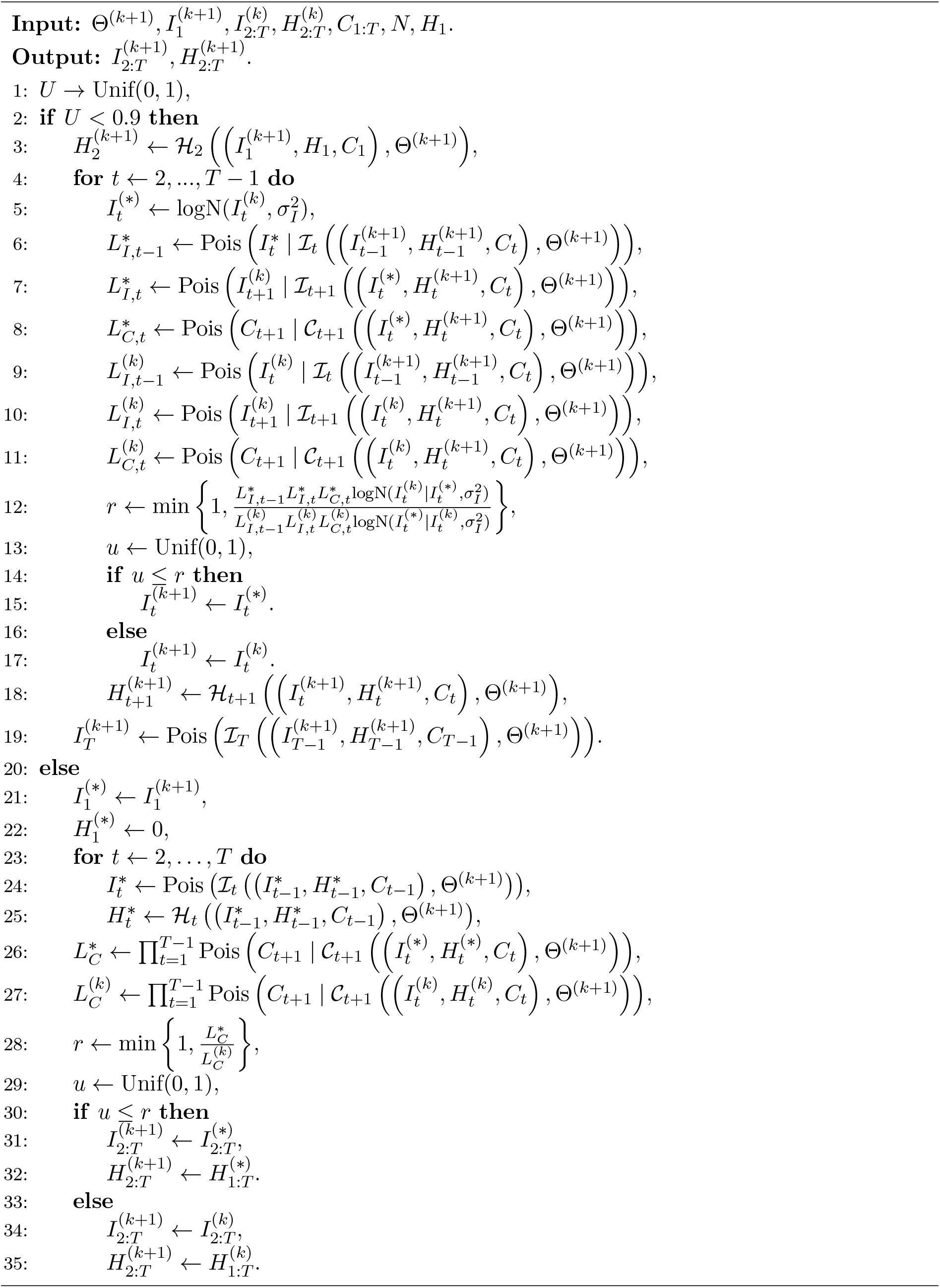

**Figure 9:**
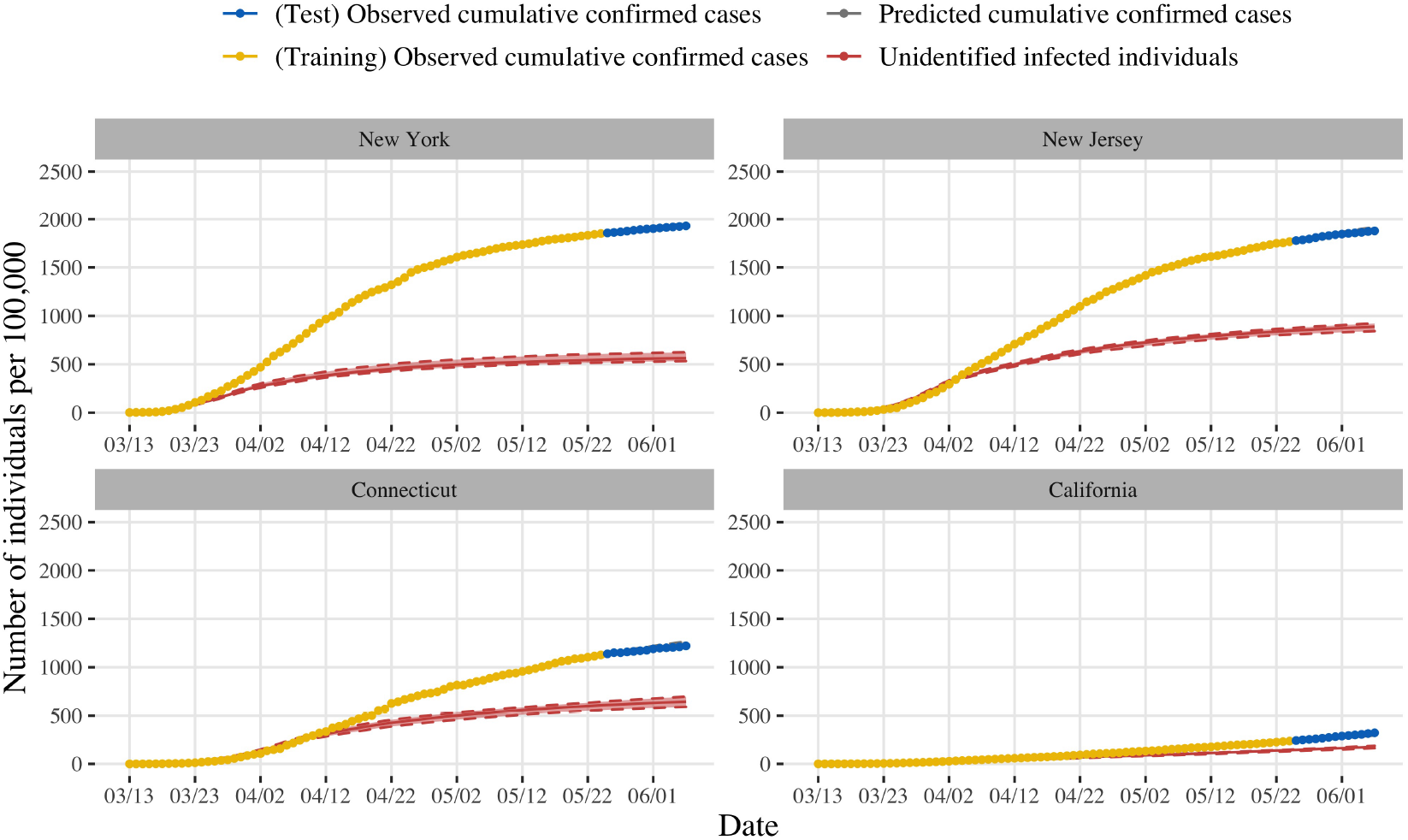
The state-specific trends of simulated confirmed cases (*C*) and unidentified infected (*I*) per 100,000 from March 13 to June 6, 2020. The points represent the observed cumulative confirmed cases.

For simulation of the second wave of the infection, we defined a “reopening” operation 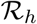 on *α*(*t*):

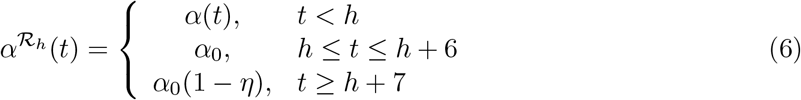

which implies the stringent interventions would be re-enforced one week after the business was reopened at time *h*.

## References

Anderson RM, Anderson B, May RM (1992). Infectious diseases of humans: dynamics and control. Oxford University Press.

Andrieu C, De Freitas N, Doucet A, Jordan MI (2003). An introduction to mcmc for machine learning. Machine Learning, 50(1-2): 5–43.

Blackwood JC, Childs LM (2018). An introduction to compartmental modeling for the budding infectious disease modeler. Letters in Biomathematics, 5(1): 195–221.

Bolstad WM, Curran JM (2016). Introduction to Bayesian statistics. John Wiley & Sons.

Centers for Disease Control and Prevention (2020). Return to work for healthcare personnel with confirmed or suspected covid-19.

Chib S, Greenberg E (1995). Understanding the metropolis-hastings algorithm. The American Statistician, 49(4): 327–335.

Chinese Center for disease control and prevention (2020). Epidemic update and risk assessment of 2019 novel coronavirus.

Ghosh JK, Delampady M, Samanta T (2006). Bayesian inference and decision theory. An Introduction to Bayesian Analysis: Theory and Methods, 29–63.

Holshue ML, DeBolt C, Lindquist S, Lofy KH, Wiesman J, Bruce H, et al. (2020). First case of 2019 novel coronavirus in the united states. New England Journal of Medicine.

Hu Z, Song C, Xu C, Jin G, Chen Y, Xu X, et al. (2020). Clinical characteristics of 24 asymptomatic infections with covid-19 screened among close contacts in nanjing, china. Science China Life Sciences, 63(5): 706–711.

Jin B (2020). Live tracker: How many coronavirus cases have been reported in each u.s. state?

Jones JH (2007). Notes on r0. California: Department of Anthropological Sciences, 323: 1–19.

Lauer SA, Grantz KH, Bi Q, Jones FK, Zheng Q, Meredith HR, et al. (2020). The incubation period of coronavirus disease 2019 (covid-19) from publicly reported confirmed cases: estimation and application. Annals of Internal Medicine, 172(9): 577–582.

Minnesota Daily (2020). The last days of may: A visual timeline of the george floyd protests.

National Health Commission of the People’s Republic of China (2020). Distribution of covid-19 cases in the world 2020.

Office of Public Affairs (2020a). Cdc confirms possible first instance of covid-19 community transmission in california.

Office of Public Affairs (2020b). Order of the state public health officer.

Office of Public Affairs (2020c). Order of the state public health officer.

Office of Public Affairs (2020d). Two confirmed cases of novel coronavirus in california.

Soetaert KE, Petzoldt T, Setzer RW (2010). Solving differential equations in r: package desolve. Journal of statistical software, 33.

Soubeyrand S (2016). Construction of semi-markov genetic-space-time seir models and inference. Journal de la Société Française de Statistique, 157(1): 129–152.

State of Connecticut Governor Ned Lamont (2020a). Governor lamont and governor raimondo to align hair salon and barber shop reopenings in early june.

State of Connecticut Governor Ned Lamont (2020b). Governor lamont announces classes at k-12 public schools will remain canceled for the rest of the academic year.

State of Connecticut Governor Ned Lamont (2020c). Governor lamont announces first positive case of novel coronavirus involving a connecticut resident.

State of Connecticut Governor Ned Lamont (2020d). Governor lamont releases rules for businesses under first phase of connecticut’s reopening plans amid covid-19.

State of Connecticut Governor Ned Lamont (2020e). Governor lamont signs executive order asking connecticut businesses and residents: ‘stay safe, stay home’.

State of Connecticut Governor Ned Lamont (2020f). Northeast governors from connecticut, new york, new jersey announce collective measures to combat spread of covid-19.

State of New Jersey Governor Phil Murphy (2020a). Governor murphy, acting governor oliver, and commissioner persichilli announce first presumptive positive case of novel coronavirus in new jersey.

State of New Jersey Governor Phil Murphy (2020b). Governor murphy and superintendent callahan announce administrative order authorizing in-person sales at car dealerships, motorcycle dealerships, boat dealerships, and bike shops.

State of New Jersey Governor Phil Murphy (2020c). Governor murphy announces statewide stay at home order, closure of all non-essential retail businesses.

State of New Jersey Governor Phil Murphy (2020d). Governor murphy, governor cuomo, governor lamont, governor carney announce multi-state agreement on beaches ahead of memorial day weekend.

State of New Jersey Governor Phil Murphy (2020e). Governor murphy signs executive order.

State of New Jersey Governor Phil Murphy (2020f). Governor murphy signs executive order permitting resumption of non-essential construction, curbside pickup at non-essential retail businesses, and gatherings in cars.

State of New Jersey Governor Phil Murphy (2020g). Governor murphy signs executive order reopening state parks and golf courses.

State of New York Governor Andrew Cuomo (2020a). Amid ongoing covid-19 pandemic, governor cuomo announces schools and college facilities statewide will remain closed for the rest of the academic year.

State of New York Governor Andrew Cuomo (2020b). Amid ongoing covid-19 pandemic, governor cuomo announces three regions of new york state ready to begin reopening may 15th.

State of New York Governor Andrew Cuomo (2020c). Governor cuomo signs the ‘new york state on pause’ executive order.

State of New York Governor Andrew Cuomo (2020d). Massachusetts joins new york, new jersey, connecticut, pennsylvania, delaware and rhode island’s multi-state council to get people back to work and restore the economy.

State of New York Governor Andrew Cuomo (2020e). Video, audio, photos & rush transcript: Amid ongoing covid-19 pandemic, governor cuomo announces seventh region hits benchmark to begin reopening tomorrow.

State of New York Governor Andrew Cuomo (2020f). Video, audio, photos & rush transcript: Amid ongoing covid-19 pandemic, governor cuomo issues executive order requiring all people in new york to wear masks or face covering in public.

State of New York Governor Andrew Cuomo (2020g). Video, audio, photos & rush transcript: During coronavirus briefing, governor cuomo issues executive order allowing state to increase hospital capacity.

Tan J, Jiang Y, Tian T, Wang X (2020). P-sihr probabilistic graphical model: an applicable model of covid-19 in estimating the number of infectious individuals. Acta Mathematicae Applicatae Sinica, 43(2): 365–382.

The Center for Systems Science and Engineering (CSSE) at Johns Hopkins University (2020). Covid-19 data repository).

The New York Times (2020). Coronavirus in the u.s.: Latest map and case count.

The State Council, The People’s Republic of China (2020a). China’s progress on business resumption on april 16.

The State Council, The People’s Republic of China (2020b). State council stresses orderly resumption of production and operation.

The United States Census Bureau (2020). Table of united states counties.

The White House (2020a). 30 days to slow the spread.

The White House (2020b). Proclamation on declaring a national emergency concerning the novel coronavirus disease (covid-19) outbreak.

US Department of Labor (2020). News release bureau of labor statistics.

World Health Organization (2020a). Coronavirus disease 2019 (covid-19): situation report, 53.

World Health Organization (2020b). Coronavirus disease 2019 (covid-19): situation report, 73.

World Health Organization (2020c). Rolling updates on coronavirus disease (covid-19).

